# DISPOSITION OF ADOLESCENTS TOWARD RECEIVING COVID-19 VACCINATIONS IN VILLAVICENCIO: MYTHS AND BELIEFS

**DOI:** 10.1101/2023.01.10.23284415

**Authors:** César García Balaguera, María Fernanda Alfonso Osorio, María Camila Pardo Garzón, María Paula Echavarría

## Abstract

Global efforts regarding the COVID-19 pandemic have been focused on preventive activities, such as vaccination, since the disease is expected to become endemic. Adolescents were among the last population groups to be included in the vaccination program in Colombia, and adequate coverage has not yet been achieved in this group and in infants. It is important to understand their motivations to improve the willingness of this population to be vaccinated. A cross-sectional study was designed via an online survey in adolescents aged 14–19 years in Villavicencio Meta after validation of the survey and informed consent. The following options were provided for the question on vaccine disposition: willing, undecided, and unwilling. We described the disposition toward receiving COVID-19 vaccine using graphs and absolute and relative frequencies based on age group. A multinomial regression model was used to assess the relationship between our predictor variables and vaccine disposition in adolescents. In this study, 288 adolescents were surveyed. The risk variables for unwillingness to be vaccinated were being male (odds ratio [OR] 2.18, 95% confidence interval [CI] 0.8–5.7, p = 0.62), belonging to low social stratum (OR 2.29, 95% CI 0.9–5.88, p = 0.19), having a monthly family income of less than 1 million Colombian pesos (250 USD) (OR 2.01, 95% CI 0.8–5.16, p = 0.19), and having basic education (OR 2.59, 95% CI 0.33– 20.14, p = 0.18). Unproven myths and beliefs exert a profound influence on adolescents, which results in an unwillingness to be vaccinated. Hence, innovative public health strategies should be designed to improve the disposition to be vaccinated in this population group.

## INTRODUCTION

With the evolution of the COVID-19 pandemic, global attention has been shifted to young people, especially those under 20 years of age, who have the lowest vaccination coverage. The disease has had a higher incidence in young populations in the last year, although lethality has not increased significantly [1]. Vaccination coverage in adolescents at the beginning of the pandemic was exceptionally low because priority was given to the older adult population and those with comorbidities. Although in high-income countries vaccination of young people and adolescents began early, this has not been the case in middle- and low-income countries. Hence, the challenge of achieving adequate coverage in these countries continues to remain [2, 3, 4, 5].

The adolescent group has been included in regular vaccination programs, especially for the prevention of human papillomavirus and tetanus. However, the COVID-19 vaccine has not been integrated into regular vaccination programs, and in many countries, coverage with these biologicals is quite low, particularly in low-income countries and populations [6,7].

The strategy of bringing the vaccination program to schools has been highly effective in improving vaccination coverage for the regular program. However, this has not been the case for vaccines against COVID-19, where negative attitudes have been evidenced because of insecurity and fear of vaccines. Furthermore, nonauthorization by parents to vaccinate their children has been a key factor [8, 9].

The side effects of COVID-19 vaccines are mild in this population. Serious adverse effects are exceedingly rare, and there are already abundant data on their safety and the risk factors associated with serious post-vaccination effects. The most common are fever, pain in the area of application, headache, and general malaise. Among those requiring hospitalization, multisystemic inflammatory syndrome, anaphylaxis, myocarditis, and pericarditis have been described, but these are rare events [10, 11, 12, 13, 14].

Parental and community decisions on the vaccination of children and adolescents have so far been hesitant, partly because the knowledge about the virus is recent. Lack of evidence on the safety of vaccines and the appearance of new variants, covariants, and strains whose behavior is still uncertain both in terms of clinical behavior and vaccine response to variants are other factors. Furthermore, there is still not much certainty about the duration of effectiveness of the vaccines and the need for boosters over time, in addition to political, economic, and cultural factors that hinder the administration of COVID-19 vaccines. These factors should be analyzed by the scientific community to design effective strategies to improve the acceptance of COVID-19 vaccination [15, 16]. This study aimed to estimate the disposition toward vaccination in the adolescent age range in Villavicencio, Meta, Colombia. The secondary objective was to examine the hypothesis that previous COVID-19 infection, socioeconomic position, mental health, social connectedness, and healthy behaviors were associated with higher or lower disposition toward the vaccine among adolescents (in this study, adolescents were aged 14–19 years).

## MATERIALS AND METHODS

A cross-sectional study was designed using a previously validated online survey, which was conducted between February and March 2022. The survey inquired about the sociodemographic condition of the adolescent, social security affiliation, educational level, and COVID-19 general aspects. Moreover, as working hypotheses, subjective experiences with the disease, experiences with vaccination and beliefs, and myths and legends about COVID-19 were explored.

The disposition of adolescents toward vaccination was assessed with one question that contained three response options: Would you get vaccinated against COVID-19 with any of the vaccines currently offered?

a. Yes (willing to) b. No (Unwilling) c. Thinking about it (Undecided) Finally, we asked about their COVID-19 vaccination status, their perception of vaccines, myths and legends about the disease and vaccines, and their willingness to be vaccinated.

The study was conducted in 10 municipalities of the city of Villavicencio. Inclusion criteria were the following: adolescents aged ≥14 years, signing an informed consent form, and answering more than 80% of the questions of the survey. Exclusion criteria were not being 14 years old, not having cognitive or motor capacity to complete the survey, and being pregnant.

The project was approved by the subcommittee on bioethics in research of the Cooperative University of Colombia, bioethics concept no. BIO254, march 1, 2022. The risk of participating in the survey was minimal according to Resolution No. 008430 of 1993, which establishes the scientific, technical, and administrative standards for health research in Colombia.

We described the disposition toward the administration of the COVID-19 vaccine using graphs as well as absolute and relative frequencies based on age group. We used a multinomial regression model to assess the relationship between our predictor variables and vaccine disposition in adolescent respondents aged 14–19 years. The results from the multinomial model were expressed as odds ratios (OR) along with 95% confidence intervals (CI) and p-values.

## RESULTS

A total of 291 people between 14 and 19 years of age were surveyed; 3 people were excluded because they did not meet the age range. The sociodemographic characteristics of the 288 participants are described in Table No. 1. It was observed that 44.83% were male, 54.83% were female, and 0.34% were others. The average age was 16.4 years; 6.9% of the respondents were 14 years old, 76.2% were 15–17 years old, and 16.9% were 18–19 years old; 97.59% were single, 1.72% were in a union, and 0.69% were married.

**Table No.1.** Sociodemographic variables

| SOCIODEMOGRAPHIC VARIABLES |  |  |  |
| --- | --- | --- | --- |
| Variable | Category | n | % |
| Age (years) | ≤14 | 20 | 6,9 |
|  | 15-17 | 221 | 76,2 |
|  | 18-19 | 49 | 16,9 |
| Sex | Female | 159 | 54,83 |
|  | Male | 130 | 44,83 |
|  | Others | 1 | 0,34 |
| Marital status | Single | 283 | 97,59 |
|  | Married | 2 | 0,69 |
|  | Free union | 5 | 1,72 |
| Housing strata | 1 | 45 | 15,52 |
|  | 2 | 66 | 22,76 |
|  | 3 | 141 | 48,62 |
|  | 4 | 27 | 9,31 |
|  | 5 | 11 | 3,79 |
| District in which they lived | 1 | 15 | 4,67 |
|  | 2 | 37 | 11,53 |
|  | 3 | 22 | 6,85 |
|  | 4 | 55 | 17,13 |
|  | 5 | 74 | 23,05 |
|  | 6 | 20 | 6,23 |
|  | 7 | 46 | 14,33 |
|  | 8 | 17 | 5,3 |
|  | 9 | 17 | 5,3 |
|  | 10 | 17 | 5,3 |
| Monthly income level | Less than \$1'000,000; less than US\$250 | 105 | 36,21 |
| | 1–3 million pesos; US\$250–750 | 132 | 45,52 |
| | 3–5 million; US\$750–1253 | 30 | 10,34 |
| | More than US\$5 million; more than US\$1253 | 23 | 7,93 |
| Educational level | School: elementary to middle school | 261 | 90 |
|  | University | 28 | 9,66 |
|  | None | 1 | 0,34 |
| Social security affiliation | Subsidized regime | 130 | 44,83 |
|  | Contributory regime | 136 | 46,9 |
|  | Special regime | 24 | 8,28 |
| Dropped out of school or work during the pandemic | Yes | 49 | 16,9 |
|  | No | 241 | 83,1 |

Source: own database

**Table No.2.**
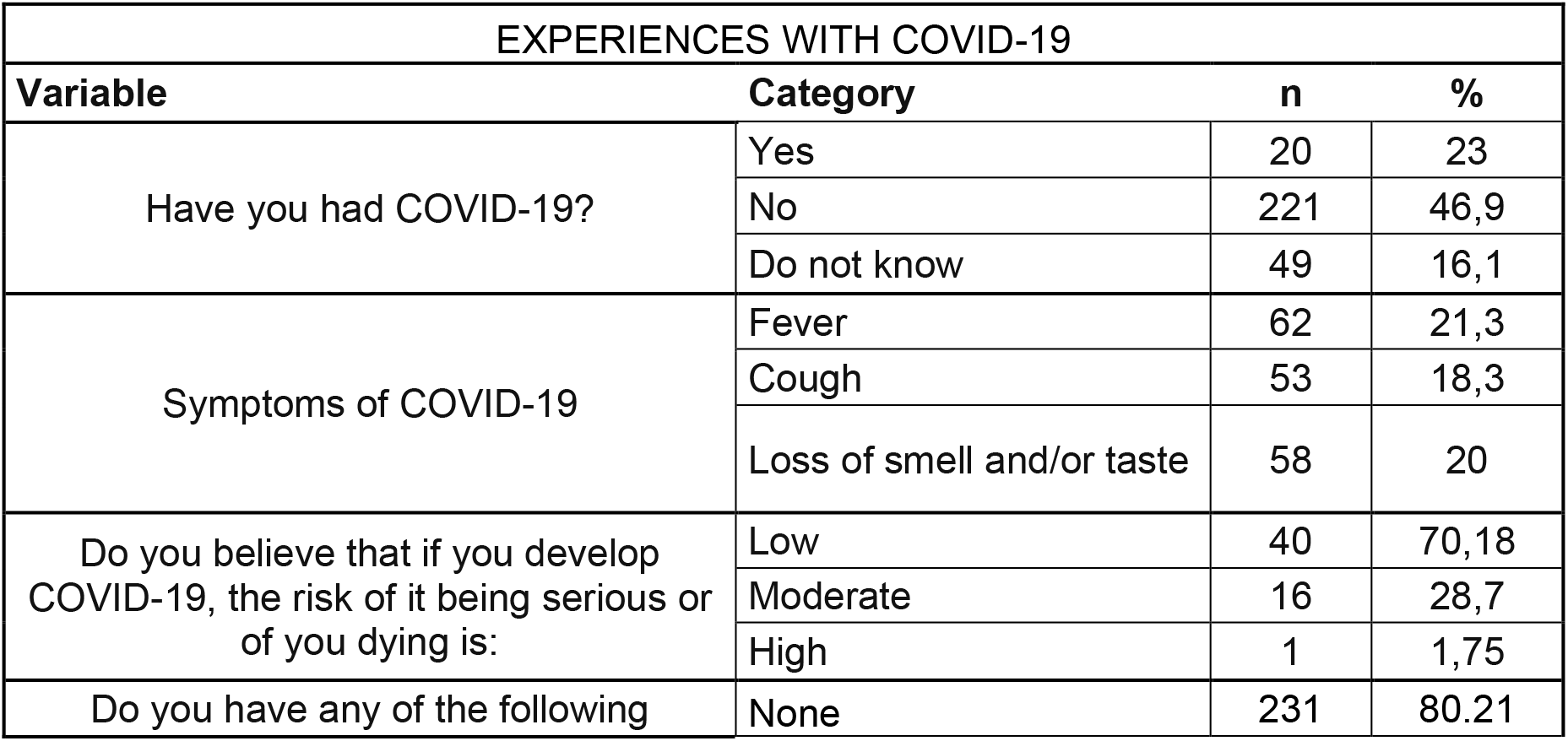
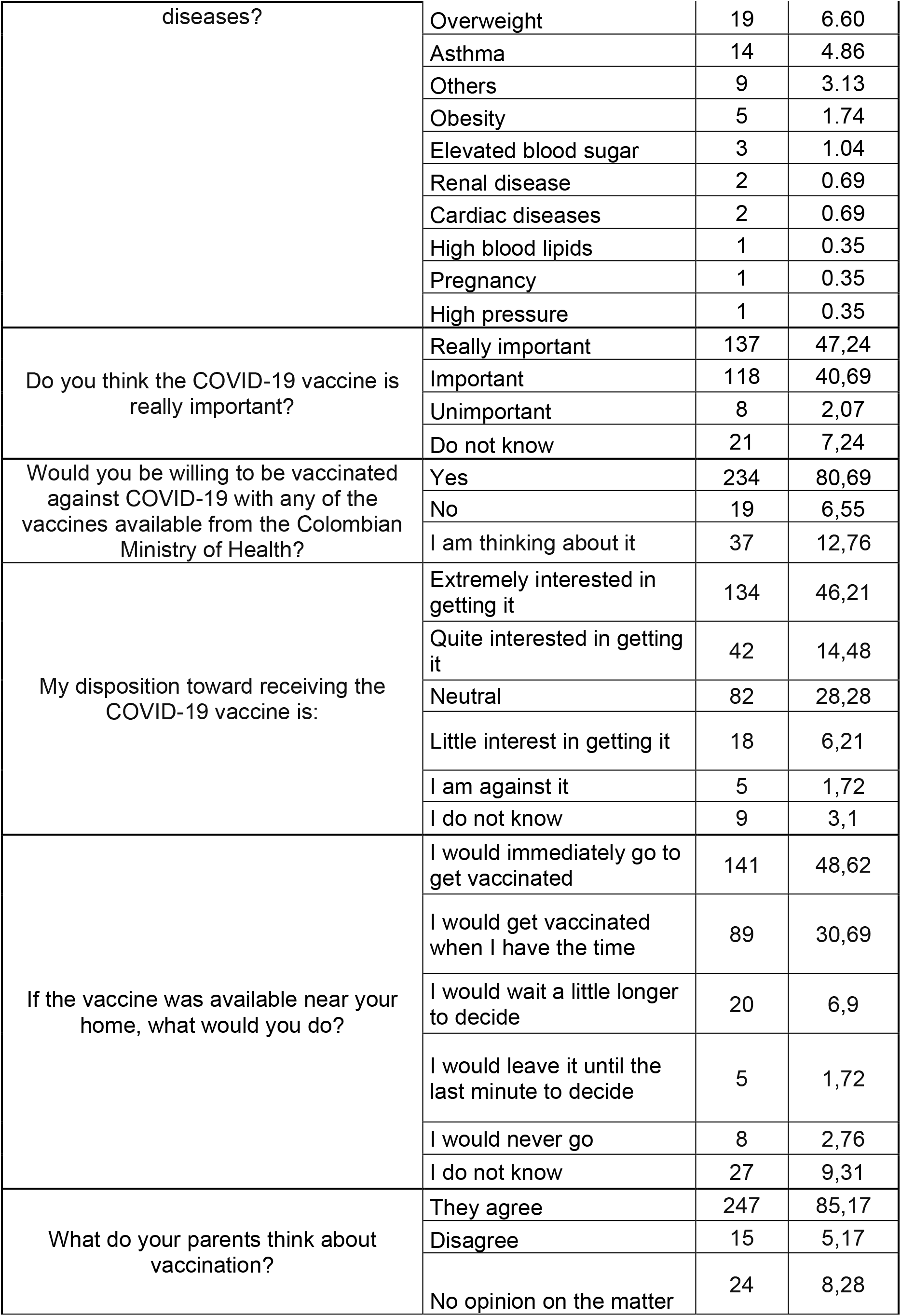
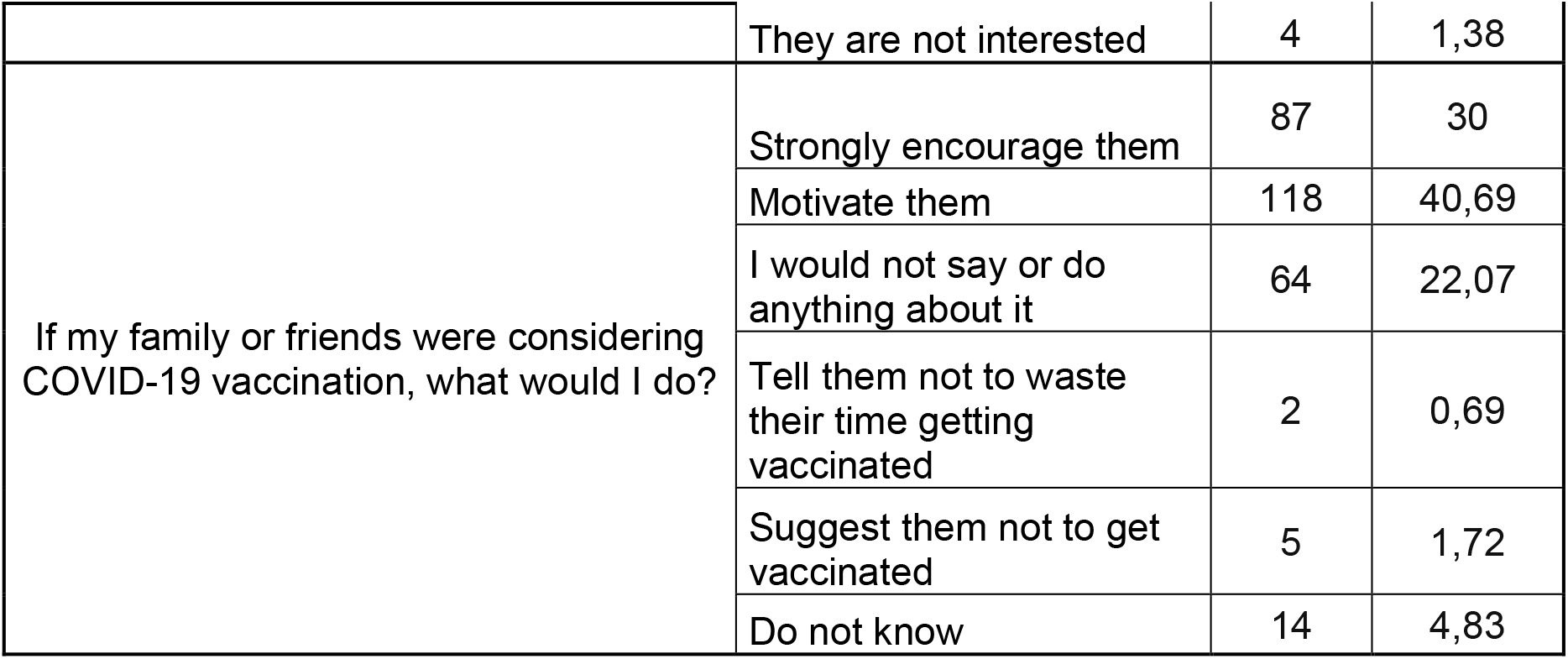
Experiences with COVID-19 and vaccination

**Table No.3.** Multinomial analysis

| DISPOSITION OF ADOLESCENTS TOWARD VACCINATION AGAINST COVID-19 |  |  |  |  |  |  |  |
| --- | --- | --- | --- | --- | --- | --- | --- |
|  |  | WOULD NOT BE VACCINATED |  |  | UNDECIDED |  |  |
| Variable | Risk factor | OR | CI | p | OR | CI | p |
| **Sociodemographic characteristics** |  |  |  |  |  |  |  |
| Age | <14 Years | 1,85 | 0,4-8,3 | 0,56 | 1,7 | 0,3-9,37 | 0,56 |
| Sex | Male | 2,18 | 0,8-5,7 | 0,19 | 2,69 | 0,85-8,49 | 0,19 |
| Social stratum | Low Stratum | 2,29 | 0,9-5,88 | 0,18 | 0,55 | 0,17-1,73 | 0,18 |
| Income level | Less than \$1 million per month | 2,01 | 0,8-5,16 | 0,19 | 0,57 | 0,18-1,75 | 0,19 |
| Educational level | Basic education (high school) | 2,59 | 0,33-20,14 | 0,18 | 0,5 | 0,03-8,91 | 0,18 |
| Affiliation to Health Social Security | Subsidized regime (low income) | 1,26 | 0,49-3,22 | 0,57 | 0,5 | 0,16-1,57 | 0,56 |
| **Experiences in the face of COVID-19** |  |  |  |  |  |  |  |
| Has had COVID-19 | No | 0,35 | 0,11-1,09 | 0,16 | 0,56 | 0,13-2,36 | 0,16 |
|  | Possible that I had it without diagnosis | 0,82 | 0,27-2,49 | 0,19 | **4,07** | **1,12-14,8** | **0,005** |
| Risk perception | Low | 0,68 | 0,14-3,33 | 0,54 | 0,48 | 0,02-8,32 | 0,9 |
|  | Intermediate | 1,73 | 0,54-5,51 | 0,54 | 1,08 | 0,28-4,18 | 0,9 |
| Comorbidities | None | 1,42 | 0,39-5,07 | 0,67 | 0,85 | 0,18-4,06 | 0,67 |
| **Beliefs, myths, and perceptions about COVID-19** |  |  |  |  |  |  |  |
| Does COVID-19 exist? | No | **7,09** | **1,62-31,02** | **0,009** | **1,5** | 0,29-7,52 | 0,57 |
| Created in a laboratory | Yes | 1,95 | 0,74-5,12 | 0,25 | 0,75 | 0,23-2,43 | 0,75 |
| Created to reduce population size | Yes | 2,14 | 0,78-5,84 | 0,96 | 0,95 | 0,28-3,16 | 0,907 |
| Created due to money and politics | Yes | **6,79** | **2,35-19,59** | **0,0005** | 2 | 0,59-6,7 | 0,12 |
| Designed for sterilization | Yes | **5,38** | **1,52-18,93** | **0,00016** | 1,33 | 0,32-5,45 | 0,13 |
| Has technologies designed to damage health | Yes | **13,27** | **4,78-36,83** | **0,0007** | **3,33** | **1,02-10,79** | **0,0007** |
| Causes health damage | Yes | 2,65 | 0,98-7,14 | 0,46 | 0,91 | 0,29-2,88 | 0,46 |
| Vaccine can lead to death | Yes | **14,92** | **5,36-41,53** | **0,00004** | **3,89** | **1,207-12,56** | **0,00002** |
| Can you get ill with COVID-19 after vaccination? | Yes | 1,314 | 0,48-3,58 | 0,13 | 1,54 | 0,47-4,99 | 0,013 |

Regarding the housing strata, with 1 being the stratum with the lowest income and 5 being the stratum with the highest income, stratum 1 accounted for 15.28%, stratum 2 for 22.92%, stratum 3 for 48.61%, stratum 4 for 9.38%, and stratum 5 for 3.82%. In terms of the income variable, minimum wage was earned by 45% of the respondents, less than one million pesos was earned by 36% of the respondents, and more than 3 million was earned by 10% of the respondents. The sole activity of 89.93% of the respondents was studying, 9.72% studied and worked, and 0.35% only worked. Furthermore, 17.01% stopped working or studying during the pandemic.

In relation to the level of studies, 89.9% received basic education, 9.72% were in the university, and 0.35% were not studying. Regarding social security affiliation, 46.88% were in the contributory system, 44.79% in the subsidized system, and 8.33% in the special system.

Persons under 17 years of age, married people, and those with low social status, low income, and low educational level were less willing to get vaccinated.

## Experiences with COVID-19

In the survey, 46.9% of the people denied having had COVID-19, 23% claimed to have had the disease and it was confirmed, and 16% could have had it but without a confirmatory test. Moreover, 63% considered that COVID-19 will not cause serious illness or death, 31% opined that it may cause moderate illness, and 6% thought that it may lead to tragic consequences.

The surveyed respondents, given their age range, reported that 80.3% did not have comorbidities, whereas 19.7% had comorbidities, such as being overweight (6%), asthma (4%), and others (3%). A total of 47% of the respondents believed that the vaccine is really important, 41% thought it is important, 3% said it is unimportant, and 7% of the participants did not know. Hence, it is vital to emphasize medical education on the importance of vaccination and break barriers of ignorance in children.

The survey revealed that the family members of 58.28% of the respondents have fallen ill due to COVID-19, and 27.24% claimed to have a family member who has been hospitalized for COVID-19 or has died from the disease.

### Experiences with vaccination

Regarding their disposition toward vaccination, 46.2% were very willing to get vaccinated, 14.5% were quite willing to be vaccinated, 3.1% were unwilling to get vaccinated, 28.3% were neutral, and 1.72% were against vaccination.

If the vaccine was available today near their home, 48.62% would go immediately to get vaccinated, 6.9% would wait a little longer to decide, 30.69% would get vaccinated when they had time, 1.72% would leave it until the last moment to decide, 2.76% would never go, and 9.31% did not know.

A total of 85.17% of the respondents’ parents agreed with vaccination, 8.28% had no opinion, 1.38% were not interested, and 1.38% expressed no interest on the subject. If family or friends were interested in getting vaccinated, 30% of the respondents would strongly encourage them, 40.69% would motivate them, 22.07% would not say or do anything, 0.69% would ask them not to waste their time getting vaccinated, 1.72% would suggest that they not get vaccinated, and 4.83% did not know what they would do.

When asked if they were vaccinated against COVID-19, 66.55% of the respondents said they have already been vaccinated, whereas 33.45% have not been vaccinated. Regarding fear of getting vaccinated, 88.97% were not afraid of vaccination, whereas 11.03% reported fear of vaccination. Of those who were not scared of vaccination, 86.55% planned to be vaccinated.

The findings showed that 97.93% of the relatives of the respondents have been vaccinated against COVID-19. Furthermore, 17.59% affirmed that being already vaccinated they will not get sick from COVID-19.

Regarding postvaccination symptoms, 35.52% reported such symptoms, the most frequent being muscle pain, fever, and chills. At the time of the survey, 51.38% had been vaccinated with Pfizer, Moderna 7.59%, Janssen 3.10%, AstraZeneca 2.07%, and Sinovac 0.69%. A total of 33.79% reported one dose and 31.03% two doses.

### Beliefs, myths, and COVID-19 perceptions

When asked if they believe that COVID really exists, 95.5% said yes. To the question, “Do you believe that the virus was created by people in a laboratory to cause harm?” 50.69% said yes. Moreover, 53.79% believed that it was created to reduce the size of the world population, 35.8% believed that the virus was created to make money and make politics, 7.24% of the respondents believed that the vaccine was created to sterilize the population, and 12.76% believed that the vaccine has some component that undermines the welfare of the individual. In addition, 21.72% believed that the vaccine could cause alterations in coagulation, heart disease, infertility, and neurological disease.

Additionally, 16.21% believed that the vaccine can cause death slowly and 62.41% believed that after vaccination they could get sick with COVID-19. To the question, “do you consider that you can stop protecting yourself with measures such as face masks and avoiding crowds after vaccination?” 14.83% said yes. Moreover, 30.34% believed they will need more than two doses of the vaccine to be protected.

### Bivariate analysis

Results of the multinomial regression model, adjusted for multiple covariates, for the sociodemographic variables suggested that the risk variables for not willing to be vaccinated included being male (OR 2.18, 95% CI 0.8–5.7, p = 0.62), belonging to a low social stratum (OR 2.29, 95% CI 0.9–5.88, p = 0.19), having a family income of less than 1 million Colombian pesos (250 USD) (OR 2.01, 95% CI 0.8–5.16, p = 0.19), and having basic education (OR 2.59, 95% CI 0.33– 20.14, p = 0.18).

Regarding experiences with COVID-19, not knowing if they have had COVID-19, although they could have had it, was a risk factor for not being vaccinated in the undecided population (OR 4.07, 95% CI 1.12–14.8, p = 0.0005). In connection to myths, beliefs, and perceptions about COVID-19 in the surveyed adolescents, the risk factors associated with not getting vaccinated were the nonexistence of COVID-19 (OR 7.09, 95% CI 1.62–31.02, p = 0.009), believing that COVID-19 was created to make money and politics (OR 6.79 95% CI 2.35–19.59, p = 0.0005), believing that the COVID-19 vaccine causes sterility (OR 5.38, 95% CI 1.52–18.93, p = 0.0016), and believing that it can cause death in the medium or long term (OR 14.92, 95% CI 5.36–41.53, p = 0.00001.

## DISCUSSION

The results indicate that most of the adolescents surveyed are willing to be vaccinated against COVID-19; however, the group of undecided and anti-vaccine respondents represents almost 20% of those surveyed. Hence, it important to identify the variables associated with this behavior. The findings of the study suggest that being less than 18 years of age, male sex, low strata, and low educational level are the sociodemographic variables most associated with being anti-vaccine or undecided. The results strongly imply that myths and beliefs have a profound influence on the low predisposition to be vaccinated in the population group of adolescents surveyed. Various myths and theories, such as the denial of the existence of COVID-19, that it was created to make money and politics, that it can cause sterility, that it contains technologies that damage health, and that it can cause death in the medium or long term, are quite accepted and are part of the motivations for being undecided about vaccination. This attitude calls for improving information, education, and communication strategies for adolescents as they are somehow very susceptible to the rumors and myths that circulate by word of mouth and social networks.

Better strategies are needed regarding informed consent. Hence, for adolescents to consent to COVID-19 vaccination, they must be able to understand the reason vaccination was recommended, including a broad explanation on the benefits of protecting themselves and others from contracting a disabling and potentially serious disease. Moreover, they need to understand in general terms the nature of the procedure; for example, that it would involve an injection in the arm that could cause pain at the injection site. In addition, healthcare professionals are required to inform those to be vaccinated of the risks for the average individual, which they are entitled to know.

We are in a transition phase of the pandemic and are moving toward an endemic phase owing to a decrease in the number of susceptible people, either because they have become ill or because of the effects of vaccination. However, the appearance of new strains, variants, and covariants implies that we must continue taking preventive measures in public health and, of course, vaccination continues to be fundamental. It is also evident that children and adolescent groups have been the last to be included in the vaccination plans and the coverage is still low. As of May 2022, in Colombia, it was below 35% for the second dose. It is, therefore, necessary to focus on the undecided and at-risk groups to reinforce vaccination efforts in these groups.

Information and communication strategies should be improved in the low stratum populations and in those with low educational level since these groups are dominated by the undecided and those not inclined to vaccination, thus generating high-risk conglomerates that need to be vaccinated.

Young and undecided schoolchildren spend a lot of time on social networks, so this channel is important in improving the knowledge and understanding about vaccination. In addition, students who have had a previously confirmed COVID-19 infection or probable infection were more prone to choose not getting vaccinated or be undecided. Therefore, better information is needed to answer any questions regarding immunity in those who have had the disease. As we learn more about the range of immune responses after exposure to the virus, this knowledge is shared with younger populations. Hence, it should be made clear that reinfection is possible. Furthermore, our study indicates that misinformation, both online and offline, has a direct impact on confidence and the intention to get vaccinated, as reported by Loomba [17]. Schools should desirably include in their curriculum the study of the history of vaccines and their profound impact on primary prevention, thereby increasing the level of knowledge and acceptance.

The impact of myths on adolescents is extremely high and greatly compromises their decisions on health and self-care [18,19]. These decisions also tend to be contrary to the opinions of adults and are, therefore, a part of their attitude toward emancipation. However, the most serious aspect is that it puts their health and, in the case of COVID-19, the health of other population groups at elevated risk [20,21,22]. Their high dependence on social networks and the digital era generates greater difficulties in controlling. Hence, it is a priority to improve the role of the state in monitoring, controlling, and supervising the quality and veracity of health information, especially against conspiracy theories and myths that surround the COVID-19 pandemic. [23,24,25]

The proportions of the population willing to be vaccinated vary significantly around the world, and cultural and social contexts are the determinants [26,27,28]. However, some common risk features are observed, such as low-income strata or educational level and living in rural areas [29,30,31]. These coincide with our findings from the present study, which suggest that intersectoral interventions are needed to achieve effective changes in the behavior of people, especially in anti-vaccine groups and in those who are undecided about being vaccinated.

The immediate future in the face of COVID-19 is uncertain. The discovery of new strains and variants, alterations in clinical behavior, limited knowledge about the duration of immunity achieved by available vaccines, anti-vaccine movements, and political and cultural contexts represent a great challenge to public health. It is important to allocate more resources to understand the perceptions of people about their health risks, generate systems of social participation and citizen action to protect their health and well-being, and strengthen primary health care. By doing so, the levels of acceptability of preventive programs and, in this case, of vaccination programs can be improved.

One of the limitations of our study is the limited sample size. As quarantine was in full force at the time of our study, access to certain population groups was overly complicated. Moreover, questions regarding family perceptions or myths are susceptible to bias, especially because we could not guarantee the confidentiality of the survey.

## Data Availability

All data are in the manuscript.

## Source of funding

Universidad Cooperativa de Colombia, Villavicencio, Meta

## Conflicts of Interest

None declared

## Notes

### Competing Interest Statement

The authors have declared no competing interest.

### Clinical Trial

This study is not a clinical trial

### Funding Statement

This research project was funded by the Universidad Cooperativa de Colombia. Support for this research was provided through the payment of research hours to the principal investigator CGB, for a value of $6,046,000 (US$1,592), which corresponds to 4 hours of research per week, for 12 months. This institution had no influence on the design, objectives, execution and conclusions of the study.

## References

1. Fazel M, Puntis S, White SR, Townsend A, Mansfield KL, Viner R, et al. Willingness of children and adolescents to have a COVID-19 vaccination: Results of a large whole schools survey in England. EClinicalMedicine [Internet]. 2021;40(101144):101144. Available from: http://dx.doi.org/10.1016/j.eclinm.2021.101144

2. Gurwitz D. COVID-19 vaccine hesitancy: Lessons from Israel. Vaccine [Internet]. 2021;39(29):3785–6. Available from: http://dx.doi.org/10.1016/j.vaccine.2021.05.085

3. Wodi AP, Murthy N, Bernstein H, McNally V, Cineas S, Ault K. Advisory Committee on Immunization Practices recommended immunization schedule for children and adolescents aged 18 Years or Younger - United States, 2022. MMWR Morb Mortal Wkly Rep [Internet]. 2022;71(7):234–7. Available from: http://dx.doi.org/10.15585/mmwr.mm7107a2

4. Ledford H. Should children get COVID vaccines? What the science says. Nature [Internet]. 2021;595(7869):638–9. Available from: http://dx.doi.org/10.1038/d41586-021-01898-9

5. Bagateli LE, Saeki EY, Fadda M, Agostoni C, Marchisio P, Milani GP. COVID-19 vaccine hesitancy among parents of children and adolescents living in Brazil. Vaccines (Basel) [Internet]. 2021;9(10):1115. Available from: http://dx.doi.org/10.3390/vaccines9101115

6. Murthy BP, Zell E, Kirtland K, Jones-Jack N, Harris L, Sprague C, et al. Impact of the COVID-19 pandemic on administration of selected routine childhood and adolescent vaccinations-10 US jurisdictions. 2020.

7. McKinnon B, Quach C, Dubé È, Tuong Nguyen C, Zinszer K. Social inequalities in COVID-19 vaccine acceptance and uptake for children and adolescents in Montreal, Canada. Vaccine [Internet]. 2021;39(49):7140–5. Available from: http://dx.doi.org/10.1016/j.vaccine.2021.10.077

8. Urrunaga-Pastor D, Herrera-Añazco P, Uyen-Cateriano A, Toro-Huamanchumo CJ, Rodriguez-Morales AJ, Hernandez AV, et al. Prevalence and factors associated with parents’ non-intention to vaccinate their children and adolescents against COVID-19 in Latin America and the Caribbean. Vaccines (Basel) [Internet]. 2021;9(11):1303. Available from: http://dx.doi.org/10.3390/vaccines9111303

9. Zychlinsky Scharff A, Paulsen M, Schaefer P, Tanisik F, Sugianto RI, Stanislawski N, et al. Students’ age and parental level of education influence COVID-19 vaccination hesitancy. Eur J Pediatr [Internet]. 2022;181(4):1757–62. Available from: http://dx.doi.org/10.1007/s00431-021-04343-1

10. Buonsenso D, Munblit D, De Rose C, Sinatti D, Ricchiuto A, Carfi A, et al. Preliminary evidence on long COVID in children. Acta Paediatr [Internet]. 2021;110(7):2208–11. Available from: http://dx.doi.org/10.1111/apa.15870

11. Osmanov IM, Spiridonova E, Bobkova P, Gamirova A, Shikhaleva A, Andreeva M, et al. Risk factors for post-COVID-19 condition in previously hospitalised children using the ISARIC Global follow-up protocol: a prospective cohort study. Eur Respir J [Internet]. 2022;59(2):2101341. Available from: http://dx.doi.org/10.1183/13993003.01341-2021

12. Wallace M, Woodworth KR, Gargano JW, Scobie HM, Blain AE, Moulia D, et al. The Advisory Committee on Immunization Practices’ interim recommendation for use of Pfizer-BioNTech COVID-19 vaccine in adolescents aged 12-15 years - United States, May 2021. MMWR Morb Mortal Wkly Rep [Internet]. 2021;70(20):749–52. Available from: http://dx.doi.org/10.15585/mmwr.mm7020e1

13. Snapiri O, Rosenberg Danziger C, Shirman N, Weissbach A, Lowenthal A, Ayalon I, et al. Transient cardiac injury in adolescents receiving the BNT162b2 mRNA COVID-19 vaccine. Pediatr Infect Dis J [Internet]. 2021;40(10):e360–3. Available from: http://dx.doi.org/10.1097/inf.0000000000003235

14. Glikman D, Stein M, Shinwell ES. Vaccinating children and adolescents against severe acute respiratory syndrome coronavirus 2 (SARS-CoV-2)-The Israeli experience. Acta Paediatr [Internet]. 2021;110(9):2496–8. Available from: http://dx.doi.org/10.1111/apa.15982

15. Verger P, Peretti-Watel P, Gagneux-Brunon A, Botelho-Nevers E, Sanchez A, Gauna F, et al. Acceptance of childhood and adolescent vaccination against COVID-19 in France: a national cross-sectional study in May 2021. Hum Vaccin Immunother [Internet]. 2021;17(12):5082–8. Available from: http://dx.doi.org/10.1080/21645515.2021.2004838

16. Morgan L, Schwartz JL, Sisti DA. COVID-19 vaccination of minors without parental consent: Respecting emerging autonomy and advancing public health: Respecting emerging autonomy and advancing public health. JAMA Pediatr [Internet]. 2021;175(10):995–6. Available from: http://dx.doi.org/10.1001/jamapediatrics.2021.1855

17. Loomba S, de Figueiredo A, Piatek SJ, de Graaf K, Larson HJ. Measuring the impact of COVID-19 vaccine misinformation on vaccination intent in the UK and USA. Nat Hum Behav [Internet]. 2021;5(3):337–48. Available from: http://dx.doi.org/10.1038/s41562-021-01056-1

18. Freeman D, Lambe S, Yu L-M, Freeman J, Chadwick A, Vaccari C, et al. Injection fears and COVID-19 vaccine hesitancy. Psychol Med [Internet]. 2021;1–11. Available from: http://dx.doi.org/10.1017/S0033291721002609

19. Bozzola E, Staiano AM, Spina G, Zamperini N, Marino F, Roversi M, et al. Social media use to improve communication on children and adolescent’s health: the role of the Italian Paediatric Society influencers. Ital J Pediatr [Internet]. 2021;47(1):171. Available from: http://dx.doi.org/10.1186/s13052-021-01111-7

20. Kebede Y, Birhanu Z, Fufa D, Yitayih Y, Abafita J, Belay A, et al. Myths, beliefs, and perceptions about COVID-19 in Ethiopia: A need to address information gaps and enable combating efforts. PLoS One [Internet]. 2020;15(11):e0243024. Available from: http://dx.doi.org/10.1371/journal.pone.0243024

21. Cui T, Yang G, Ji L, Zhu L, Zhen S, Shi N, et al. Chinese residents’ perceptions of COVID-19 during the pandemic: Online cross-sectional survey study. J Med Internet Res [Internet]. 2020;22(11):e21672. Available from: http://dx.doi.org/10.2196/21672

22. Tesfaw A, Arage G, Teshome F, Taklual W, Seid T, Belay E, et al. Community risk perception and barriers for the practice of COVID-19 prevention measures in Northwest Ethiopia: A qualitative study. PLoS One [Internet]. 2021;16(9):e0257897. Available from: http://dx.doi.org/10.1371/journal.pone.0257897

23. Kricorian K, Civen R, Equils O. COVID-19 vaccine hesitancy: misinformation and perceptions of vaccine safety. Hum Vaccin Immunother [Internet]. 2022;18(1):1950504. Available from: http://dx.doi.org/10.1080/21645515.2021.1950504

24. Ullah I, Khan KS, Tahir MJ, Ahmed A, Harapan H. Myths and conspiracy theories on vaccines and COVID-19: Potential effect on global vaccine refusals. Vacunas (Engl Ed) [Internet]. 2021;22(2):93–7. Available from: http://dx.doi.org/10.1016/j.vacune.2021.01.009

25. Sallam M, Dababseh D, Eid H, Al-Mahzoum K, Al-Haidar A, Taim D, et al. High rates of COVID-19 vaccine hesitancy and its association with conspiracy beliefs: A study in Jordan and Kuwait among other Arab countries. Vaccines (Basel) [Internet]. 2021;9(1):42. Available from: http://dx.doi.org/10.3390/vaccines9010042

26. Sallam M, Dababseh D, Eid H, Hasan H, Taim D, Al-Mahzoum K, et al. Low COVID-19 vaccine acceptance is correlated with conspiracy beliefs among university students in Jordan. Int J Environ Res Public Health [Internet]. 2021;18(5). Available from: http://dx.doi.org/10.3390/ijerph18052407

27. Eberhardt J, Ling J. Predicting COVID-19 vaccination intention using protection motivation theory and conspiracy beliefs. Vaccine [Internet]. 2021;39(42):6269–75. Available from: http://dx.doi.org/10.1016/j.vaccine.2021.09.010

28. Mohamad O, Zamlout A, AlKhoury N, Mazloum AA, Alsalkini M, Shaaban R. Factors associated with the intention of Syrian adult population to accept COVID19 vaccination: a cross-sectional study. BMC Public Health [Internet]. 2021;21(1):1310. Available from: http://dx.doi.org/10.1186/s12889-021-11361-z

29. Hong J, Xu X-W, Yang J, Zheng J, Dai S-M, Zhou J, et al. Knowledge about, attitude and acceptance towards, and predictors of intention to receive the COVID-19 vaccine among cancer patients in Eastern China: A cross-sectional survey. J Integr Med [Internet]. 2022;20(1):34–44. Available from: http://dx.doi.org/10.1016/j.joim.2021.10.004

30. Jain L, Vij J, Satapathy P, Chakrapani V, Patro B, Kar SS, et al. Factors influencing COVID-19 vaccination intentions among college students: A cross-sectional study in India. Front Public Health [Internet]. 2021;9:735902. Available from: http://dx.doi.org/10.3389/fpubh.2021.735902

31. Malik AA, McFadden SM, Elharake J, Omer SB. Determinants of COVID-19 vaccine acceptance in the US. EClinicalMedicine [Internet]. 2020;26(100495):100495. Available from: http://dx.doi.org/10.1016/j.eclinm.2020.100495

32. Lazarus JV, Ratzan SC, Palayew A, Gostin LO, Larson HJ, Rabin K, et al. Author Correction: A global survey of potential acceptance of a COVID-19 vaccine. Nat Med [Internet]. 2021;27(2):354. Available from: http://dx.doi.org/10.1038/s41591-020-01226-0

33. Rhodes A, Hoq M, Measey M-A, Danchin M. Intention to vaccinate against COVID-19 in Australia. Lancet Infect Dis [Internet]. 2021;21(5):e110. Available from: http://dx.doi.org/10.1016/S1473-3099(20)30724-6

34. Harapan H, Wagner AL, Yufika A, Winardi W, Anwar S, Gan AK, et al. Acceptance of a COVID-19 vaccine in southeast Asia: A cross-sectional study in Indonesia. Front Public Health [Internet]. 2020;8:381. Available from: http://dx.doi.org/10.3389/fpubh.2020.00381

